# Ischemic Heart Disease and Cognitive Prognosis in the First Year after Stroke

**DOI:** 10.1101/2020.05.18.20106088

**Authors:** Michael J O’Sullivan, Paul Wright

**Affiliations:** UQ Centre for Clinical Research, University of Queensland, Brisbane, Australia; Department of Neurology, Royal Brisbane and Women’s Hospital, Herston, Australia; Department of Neuroimaging, Institute of Psychiatry Psychology and Neuroscience, King’s College London, UK

## Abstract

**Importance:** Cognitive impairment is the greatest single source of unmet need identified by stroke survivors. Knowledge of the factors that influence cognitive prognosis will lead to better preventive and rehabilitation strategies.

**Objective:** To identify the factors that influence general cognitive function, memory and executive function in the first year after ischemic stroke.

**Design:** Single centre longitudinal observational study.

**Setting:** Hospital stroke service.

**Participants:** A cohort of 179 patients identified within 7 days of first symptomatic ischaemic stroke were enrolled into a longitudinal cognitive study, STRATEGIC. General cognitive function, episodic memory and executive function were assessed in the first three months and again at one year after stroke. Lesion topography was defined by imaging (n=152) performed in the acute period. Cognitive evaluation was repeated at one year in 141 participants.

**Main Outcome Measures:** Montreal Cognitive Assessment (MoCA) score at 1 year. Verbal free recall (Free and Cued Selective Reminding Test) and Digit Symbol Substitution Score provided secondary outcome measures of episodic memory and executive function respectively.

**Results:** At 50±19 days after stroke, diabetes mellitus and smoking were associated with MoCA score independent of other risk and demographic factors. Lesion vascular territory was independently associated with memory while white matter lesion burden was associated with executive function. In contrast to other risk factors, ischaemic heart disease was associated with *change* in cognitive scores and MoCA score at one year but not MoCA score at 3 months. IHD was the only factor significantly associated with change over time. This association was significant independent of other factors.

**Conclusions and Relevance:** Associations between post-stroke cognition – and age, diabetes, smoking and white matter lesions – are likely to reflect the general effects of these factors on brain structure and function. These risk factors are not associated with change in cognitive function between 3 months and one year. In contrast, pre-existing ischemic heart disease was associated specifically with change in cognition over time. On average, patients with IHD showed decline in MoCA scores between 3 and 12 months while those free of IHD showed improvement. Intervention for IHD, alongside best-care stroke rehabilitation, merits investigation as a strategy to improve cognitive prognosis after stroke.

**Key Points:** *Question:* What factors predict cognitive prognosis after stroke?

*Findings:* In this hospital-based cohort, diabetes mellitus and smoking were associated with Montreal Cognitive Assessment scores at both 3 months and 1 year after first symptomatic ischemic stroke. Ischemic heart disease (IHD), in contrast to other factors, was associated with change in cognitive scores. Participants with a history of IHD showed worsening cognition on average; those free of IHD improved.

*Meaning:* Multiple factors contribute to cognitive function after stroke. IHD is associated with change between 3 and 12 months. The nexus between heart and brain in neurologic recovery requires further exploration.

## Introduction

Cognitive impairment is a common sequel of stroke^1^. Difficulties with memory and concentration are frequently reported by stroke survivors (43% and 45% respectively) and are more likely to be reported as unmet needs than other long-term complaints such as reduced mobility or speech difficulty^2^. Early neuropsychological evaluation in one cohort showed that over a quarter of patients were impaired in executive functioning, abstract reasoning, language or verbal memory^3^. In a proportion of patients, cognitive impairment goes on to become a cornerstone of post-stroke dementia^4^. At the same time, a number of studies have reported spontaneous improvement in cognition over time^5,6^.

Age, hyperglycemia, infection and early seizures have been identified as factors associated with global cognition in the first 3 months^7^. Other reports describe associations with gender, depression and previous history of stroke^6^. However, previous studies vary in the breadth of potential confounding factors that are included. A particular problem is that more comprehensive studies of risk and demographic factors were often not comprehensive in characterising stroke lesions by imaging, coding lesion location by only hemisphere, lobe or involvement of cortex^7^. Conversely, studies that have described a relationship with infarct volume or location have been unable to affirm these relationships once other factors are considered, suggesting that associations are confounded by underlying risk factors^3^. Few, if any studies, have investigated risk factors and lesion location in parallel to define independent associations in multivariable models. A related problem is that the highly variable location of stroke is linked to individual heterogeneity in post-stroke cognitive impairment, which can affect multiple domains, so that no single outcome measure provides a full understanding of the interaction of location and other factors^1^. The correlates of memory, executive function and global cognition are not likely to overlap perfectly. Another challenge is that a number of risk factors for stroke are also associated with progressive damage to brain structure by mechanisms other than focal infarction. This makes it important to differentiate between factors associated with post-stroke cognition in cross-sectional and longitudinal analyses.

The STRATEGIC study was designed to fill these gaps in current knowledge: investigation of clinical, risk and demographic factors was accompanied by delineation of lesions on imaging; associations were assessed for global cognition and two key domains, episodic memory and executive function; and were assessed both cross-sectionally within 3 months of stroke, and for change of cognitive function between 3 and 12 months. The Montreal Cognitive Assessment (MoCA) ^8^ was used to index global cognition as it is more sensitive than the MMSE to cognitive impairment in stroke survivors ^9^. Lesion location was investigated with complementary methods applied to images acquired in the acute phase: location coded radiologically by arterial territory; and by lesion delineation and mapping to standard atlases of grey and white matter regions.

## Methods

### Design and Participants

Patients with first symptomatic ischaemic stroke were recruited within 7 days of onset from a single London hyperacute stroke unit (King’s College Hospital). Inclusion criteria were: age over 50; clinical, supratentorial stroke corroborated by CT or MRI. Exclusion criteria were: previous large-artery infarct (clinically or on imaging); existing diagnosis of dementia; lack of fluency in English; active malignancy; major neurological or psychiatric disease (as defined by DSM-IV-TR); previous moderate to severe head injury (Mayo clinic classification of severity); lack of capacity to consent. Patients with impairments likely to interfere with cognitive testing, such as severe visual impairment, dyslexia or aphasia were also excluded. Consecutive patients meeting study criteria were invited to take part. All participants gave written, informed consent at enrolment. The study was approved by the London and Bromley Research Ethics Committee (ref: 13-LO-1745). The flowchart for recruitment and assessment is shown in eFigure 1.

### Stroke subtyping, risk factors and infarct delineation

At the baseline visit, clinical investigation results and risk factor status were reviewed and etiological subtypes of stroke verified or updated. Atrial fibrillation (AF) was defined as >7 mins AF measured by either admission ECG or subsequent prolonged ECG recordings. Patients were defined as having carotid stenosis if greater than 50% vessel narrowing was reported by either carotid Doppler ultrasound or CT angiography. Ischaemic heart disease (IHD) was defined by a history of either an acute coronary syndrome or a coronary artery intervention. Smoking was defined by self-report. Hypertension was identified by asking patients whether they had been diagnosed with or prescribed medication for high blood pressure. Each of these risk factors was coded as a binary variable. Clinical imaging was reviewed and infarcts classified by hemisphere and arterial territory. Infarcts involving cortex were classified into anterior (ACA), middle (MCA), or posterior (PCA) cerebral artery territories. The MCA territory was further subdivided into anterior, posterior or deep (striatocapsular territory). Non-thalamic lacunar lesions were defined as lesions in the white matter or deep grey matter with a diameter less than 15mm. These procedures led to a classification of infarcts into one of 7 categories based on location: non-thalamic lacunar infarct; thalamic infarct; and 5 classes of large artery infarct (MCA-anterior, MCA-posterior, MCA-striatocapsular, ACA, PCA). Severity of white matter disease on clinical images was graded by a consultant neurologist (MJO) using a simplified Fazekas scale ^10^. Lesion outlines were defined in 152 participants, 85 on CT and 67 on MRI. Images were acquired at a mean of 6 days after stroke onset (range 1-91 days). Lesion volume was estimated based on clinical CT and MR images by a single rater (PW) marking lesion voxels using semi-automated contouring in Jim 8 (Xynapse Systems, Essex, UK).

### Cognitive Evaluation

Six tests was performed at the baseline visit: the National Adult Reading Test Revised (NART-R)^11^, to estimate premorbid intellectual function; Montreal Cognitive Assessment (MoCA) ^8^; Digit Symbol Substitution Test (DSST), a measure of information processing speed and executive function ^12^; Trail-making Test; Digit Span, an index of working memory ^12^; and The Free and Cued Selective Reminding Task (FCSRT), a measure of verbal recall ^13^. The Montreal Cognitive Assessment (MoCA) was used as a test of general cognitive ability. The MoCA is a short test of multiple cognitive skills (e.g. attention, language and memory) designed to achieve a rating of general cognition comparable to the Mini Mental State Examination (MMSE) but with greater sensitivity to attentional and executive function ^8^.

At the one-year follow-up visit, a full medical history was taken. Any new medical events were noted and patients with a history of recurrent stroke were excluded. The MoCA, FCSRT and Digit Span tests were repeated. For MoCA and FCSRT, patients were presented with a different version of the test from the baseline visit. Choice of baseline version was counterbalanced across patients to avoid systematic bias in scores at each time point.

### Analysis and Statistics

Statistical analyses were performed using SPSS version 25 (IBM Corp. Armonk, NY). Exploratory analyses were performed using individual variables to identify those affecting cognitive scores at baseline and follow up and the difference between baseline and follow up. Hypertension, diabetes mellitus, AF, carotid stenosis, IHD, and current smoking were coded as binary variables, lesion vascular territory as a categorical variable and Fazekas score as an ordinal variable. Age, years of education, and interval between stroke onset and cognitive testing were modelled as continuous variables. Associations of change in cognition were tested in two ways, first using the numerical difference between the two scores, and second using ANCOVA with follow up score as dependent variable and baseline score as a covariate^14^. Follow up analysis used a general linear model combining candidate variables to identify which exerted independent effects on recovery or decline.

### Data Availability Statement

All data is available for sharing with qualified investigators and will be shared, in anonymized form, upon request, in accordance with the terms of the study ethical approvals. This study is registered with ClinicalTrials.gov at https://clinicaltrials.gov/show/NCT03982147.

## Results

### Demographics, Risk Factors and Subtypes of Stroke

Demographics, prevalence of cardiovascular risk factors and data on stroke subtype and vascular territory are provided in Table 1. There were 94 right hemisphere and 85 left hemisphere infarcts. Baseline cognitive evaluation took place at a median 43.5 days after stroke (IQR 33, range 22–109 days). Follow up evaluation took place at a median of 384 days after stroke (IQR 41, range 355–657 days). One patient died before the 1-year follow-up and two others had recurrent symptomatic stroke, so were excluded. Thirty-five participants either declined or did not respond to the invitation for follow-up evaluation. Follow-up data were therefore acquired in 141 of the original sample.

**Table 1.** Participant demographics, infarct locations and risk factors

| Factor |  | Baseline and<br>Follow Up | Baseline<br>Only | Statistic | p |
| --- | --- | --- | --- | --- | --- |
| n |  | 141 | 38 |  | NS |
| Age, mean (SD) | | 70.0 (9.5) | 69.5 (9.0) | $t(177) = 0.3$ | NS |
| Education, mean (SD) | | 13.4 (3.7) | 12.3 (3.3) | $t(170) = 1.8$ | NS |
| Female, n (%) | | 52 (36.9) | 8 (21.1) | $\chi^2(1, N=179) = 3.4$ | NS |
| Handedness | Right | 132 | 37 | $\chi^2(1, N=179) = 0.8$ | NS |
|  | Left | 9 | 1 |  |  |
| Ethnicity | White | 122 | 29 | $\chi^2(4, N=179) = 6.3$ | NS |
|  | Non white | 19 | 9 |  |  |
| Vascular territory | Left ACA | 2 | 0 | $\chi^2(6, N=179) = 6.4$ | NS |
|  | Left MCA | 39 | 8 |  |  |
|  | Right MCA | 45 | 13 |  |  |
|  | Left PCA | 7 | 2 |  |  |
|  | Right PCA | 10 | 6 |  |  |
|  | Left lacunar | 19 | 2 |  |  |
|  | Right lacunar | 10 | 4 |  |  |
|  | Left thalamic | 4 | 2 |  |  |
|  | Right thalamic | 5 | 1 |  |  |
| Atrial fibrillation | No | 104 | 29 | $\chi^2(1, N=179) = 0.1$ | NS |
|  | Yes | 37 | 9 |  |  |
| Carotid stenosis (>50%) | No | 125 | 32 | $\chi^2(1, N=179) = 0.6$ | NS |
|  | Yes | 16 | 6 |  |  |
| Diabetes mellitus | No | 112 | 29 | $\chi^2(1, N=179) = 0.2$ | NS |
|  | Yes | 29 | 9 |  |  |
| Hypertension | No | 50 | 10 | $\chi^2(1, N=179) = 1.1$ | NS |
|  | Yes | 91 | 28 |  |  |
| Ischaemic heart disease | No | 110 | 29 | $\chi^2(3, N=179) = 0.1$ | NS |
|  | Yes | 31 | 9 |  |  |
| Smoking at stroke | Never | 74 | 11 | $\chi^2(3, N=179) = 10.8$ | **0.01** |
|  | Quit | 43 | 14 |  |  |
|  | Current | 24 | 13 |  |  |
| Fazekas score | 0 | 12 | 4 | $\chi^2(3, N=179) = 0.5$ | NS |
|  | 1 | 57 | 14 |  |  |
|  | 2 | 46 | 14 |  |  |
|  | 3 | 26 | 6 |  |  |

Statistics are for differences between basic and extra test groups. ACA: anterior cerebral artery.
MCA: middle cerebral artery. PCA: posterior cerebral artery.

### Cross-sectional Associations with Cognition

Patients’ baseline MoCA scores were broadly normally distributed and ranged from 18 to 30 out of 30, with 60 out of 141 patients scoring below 26, the nominal threshold to be considered impaired (Figure 1A) ^8^. Hypertension, diabetes mellitus (DM) and white matter lesion load (Fazekas score) were associated with MoCA scores in the first three months after stroke. For hypertension, t(139) = 3.07, p = .003; for DM, baseline t(139) = 2.50, p = .014. MoCA scores showed a monotonic decrement with each tier of white matter lesion load (baseline F(3,137) = 4.21, p = .007). Ischemic heart disease was not associated with MoCA score in the first 3 months.

**Figure 1.**
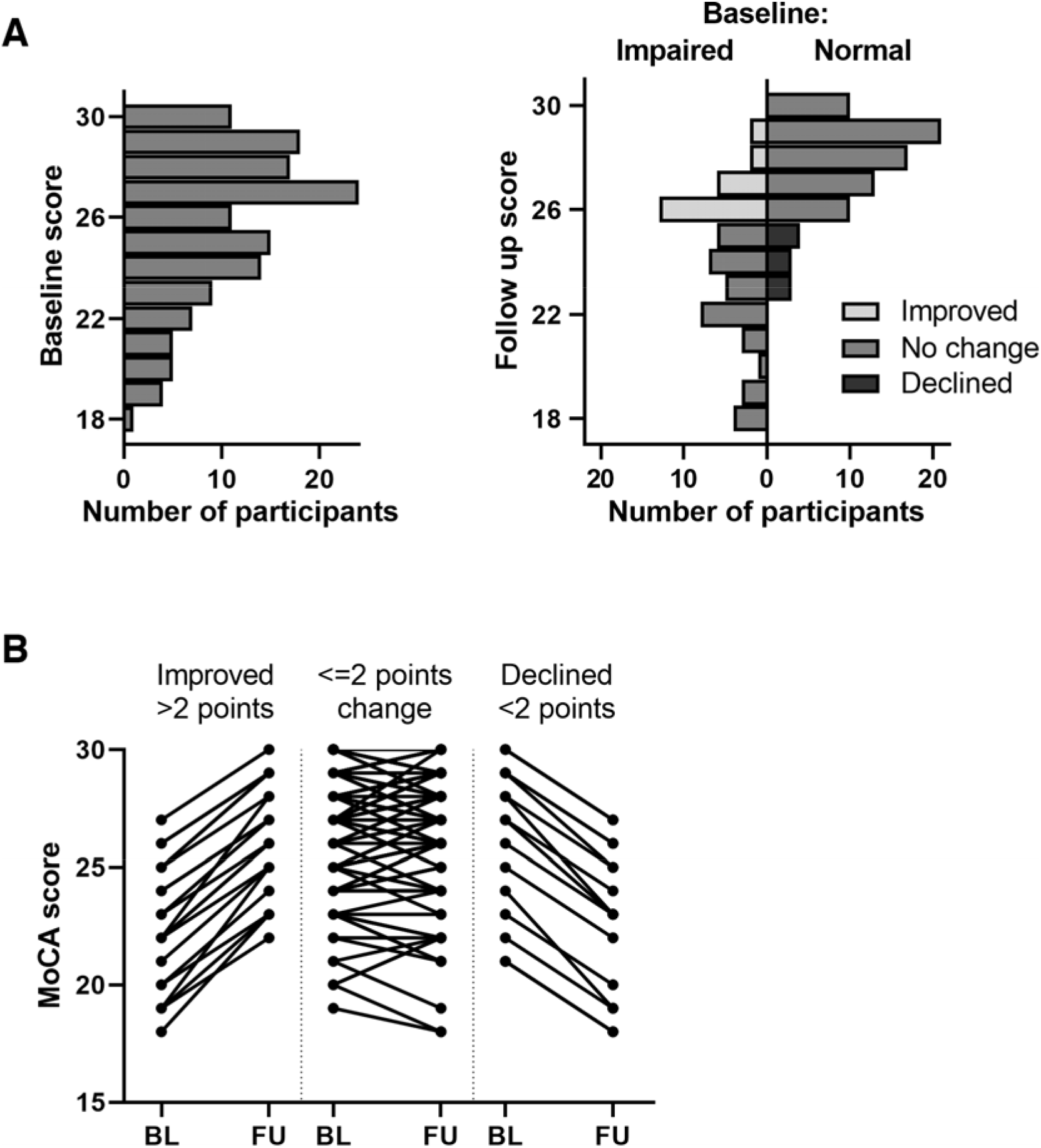
Distribution of MoCA scores and change over time.

To assess associations independent of other variables, multivariable ANCOVA models were constructed with the following variables: age; education; lesion vascular territory; smoking status; hypertension; DM; atrial fibrillation; ischemic heart disease; carotid stenosis; and Fazekas grade. The same variables were included in models for MoCA, verbal memory and executive function (Table 2). Smoking had a significant association with all cognitive scores in multivariable models. DM was associated with MoCA score and executive function. Lesion vascular territory was associated specifically with memory; white matter lesion load was associated specifically with executive function.

**Table 2.** Predictors of memory, executive function and global cognition at baseline assessment (n=179)

| Model factor | Statistic | Verbal<br>memory<br>(FCSRT free<br>recall) | Executive<br>function<br>(DSST) | MoCA |
| --- | --- | --- | --- | --- |
| ***Vascular disease pattern*** |  |  |  |  |
| Infarct location (territory) | *F*(6) | **4.48** | 1.53 | 1.06 |
|  | *p* | **<.001** | .172 | .386 |
| Infarct hemisphere | *F*(1) | 3.73 | 0.01 | 1.63 |
|  | *p* | .055 | .907 | .204 |
| Infarct hemisphere by location<br>interaction | *F*(5) | **2.29** | 1.73 | 1.98 |
|  | *p* | **.049** | .131 | .085 |
| Fazekas score | *F*(3) | 0.69 | **3.59** | 2.31 |
|  | *p* | .560 | **.015** | .078 |
| ***Risk factors*** |  |  |  |  |
| Atrial fibrillation | *F*(1) | 3.51 | 1.77 | 0.81 |
|  | *p* | .063 | .185 | .369 |
| Hypertension | *F*(1) | 0.03 | 1.29 | 0.04 |
|  | *p* | .866 | .258 | .839 |
| Diabetes mellitus | *F*(1) | 2.14 | **5.89** | **7.82** |
|  | *p* | .145 | **.016** | **.006** |
| Ischaemic heart disease | *F*(1) | 0.08 | 0.00 | 2.11 |
|  | *p* | .773 | .951 | .148 |
| Carotid stenosis | *F*(1) | 1.28 | 0.14 | 0.01 |
|  | *p* | .260 | .711 | .924 |
| Current smoker | *F*(1) | **5.35** | **10.04** | **5.79** |
|  | *p* | **.022** | **.002** | **.017** |

| ***Demographic factors*** |  |  |  |  |
| --- | --- | --- | --- | --- |
| Age | *F*(1) | **5.78** | **6.95** | 2.13 |
|  | *p* | **.017** | **.009** | .147 |
| Education | *F*(1) | 2.59 | 3.16 | **4.73** |
|  | *p* | .109 | .078 | **.031** |
| ***Overall*** |  |  |  |  |
| Overall model (corrected) | *F*(23) | **2.74** | **3.64** | **2.32** |
|  | *p* | **<.001** | **<.001** | **.001** |

Significant associations are shown in bold ( $p < 0.05$ ). FCSRT, Free and Cued Selective Reminding Test; DSST, Digit Symbol Substitution Test; MoCA, Montreal Cognitive Assessment

### Longitudinal Change in Cognitive Scores

Both improvement and deterioration in cognitive scores were observed at follow-up. For the MoCA, the majority of patients (104 or 74%) had scores within 2 points of their baseline score. Scores improved >2 points in 21 (15%) and declined >2 points in 16 (11%). Based on a MoCA cut-off score of 26, a number of patients crossed from impaired to normal and vice versa (Figure 2B). Individual trajectories are shown in Figure 2C. Neither age nor education correlated with change in score. Hypertension, DM and Fazekas score were not associated with change in cognitive scores.

Ischemic heart disease, in contrast to other risk factors, had a significant effect on MoCA score at follow up (t(139) = 2.18, p = .031) but not at baseline. Ischemic heart disease was also associated with change in MoCA score over one year, tested both as a t-test on difference scores (*t*(139) = 2.56, p = .011) and ANCOVA with baseline score as a covariate (F(1,138) = 8.4, p = .004, overall R^2^ = .545). Participants free of IHD showed a mean improvement in MoCA score over one year, while patients with IHD showed a mean deterioration (Figure 2).

**Figure 2.**
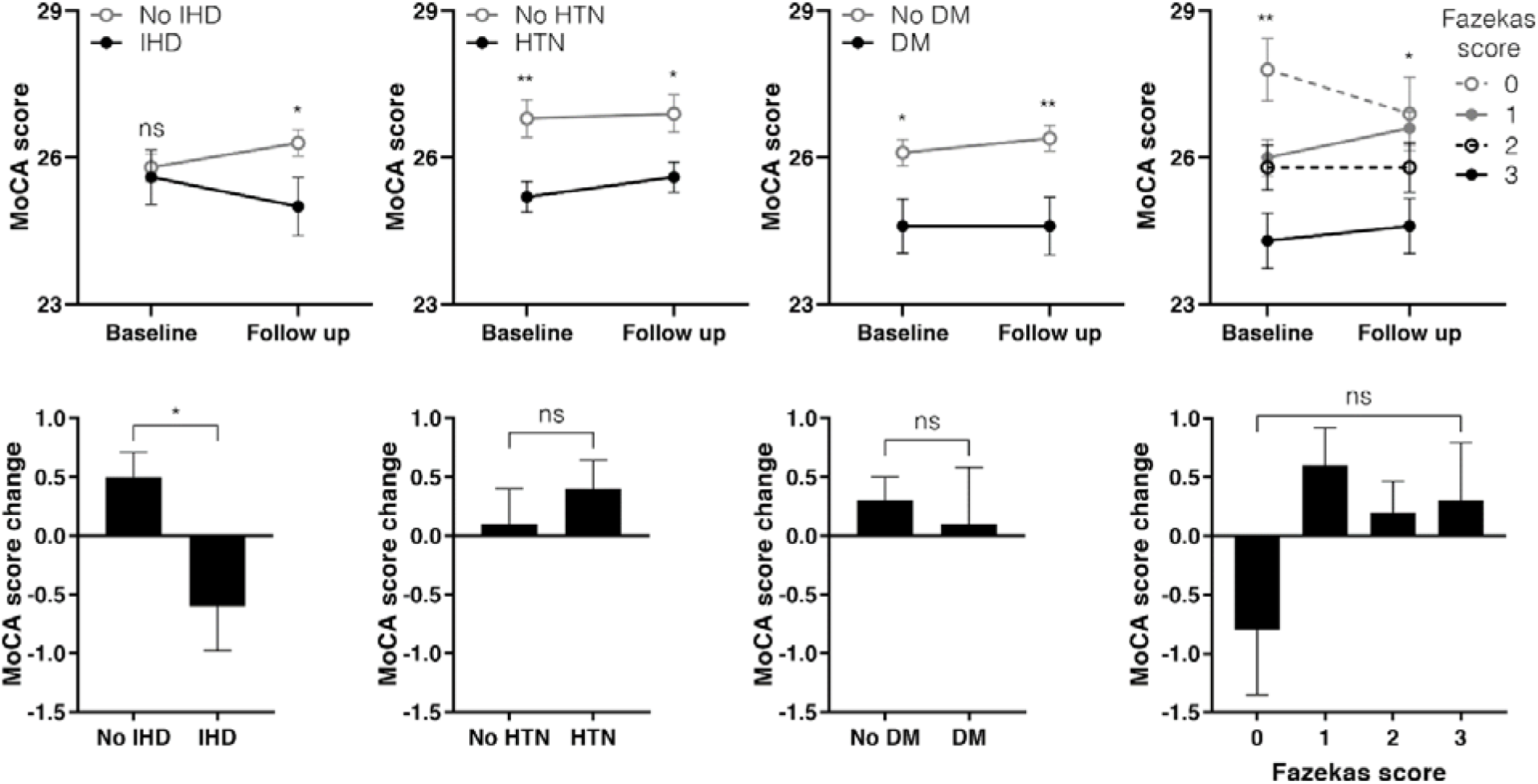
Risk factors affecting MoCA score.

A combined ANCOVA was defined using MoCA follow up score as dependent variable and with baseline MoCA score and other relevant covariates (those showing an association at either time point, namely: IHD, hypertension, DM, Fazekas score, age and education, Table 3). The model explained over half the variance (R^2^ = .570; adjusted R^2^ = .540). IHD was found to have a significant association with change in cognition, independent of other variables (F(1,131) = 8.18, p = .005). DM also had a significant effect (F(1,13) = 4.72, p = .032) in this model.

### Lesion location and Cognitive Prognosis

Inspection of memory scores for infarction in different vascular territories suggested that damage in the left thalamus and left PCA territory were responsible for the association with poor verbal recall (Figure e2). Atlas-based analysis of acute lesions suggested 4 sites of injury associated with verbal recall, 3 of which are within left PCA territory: left hippocampus (z = −3.24); left thalamus (z = −3.75); left cingulum (z = −3.32); and left inferior occipito-frontal fasciculus (z = −3.54). There was no evidence of a relationship between lesion location and change in cognitive scores. Memory scores remained poor at one year in those with left thalamic or left PCA infarction.

## Discussion

A number of factors were associated with cognitive performance after stroke, including risk factors such as smoking and diabetes mellitus, which were associated with MoCA scores independently of other factors. Lesion location had a domain-specific effect, limited to verbal memory. Consistent with previous studies, in the 9-month period after initial assessment there was evidence of both improvement and deterioration of cognitive scores for individual participants. Unlike other risk factors, IHD was associated with change in cognitive score and was a predictor of cognitive status at one year independent of status at 3 months and other risk and demographic factors. Patients with IHD showed decline, on average, while those free of IHD showed evidence of improvement.

The pattern of association with vascular risk factors is consistent with previous studies of post-stroke cognitive impairment. The Tel Aviv Brain Acute Stroke Cohort (TABASCO) study has shown that type 2 diabetes mellitus is associated with cognitive status 24 months after stroke^15^.

Furthermore, in TABASCO, type 2 diabetes mellitus was found to be associated with baseline measures of brain structure outside of the index lesion^15^. This finding suggests that diabetes has a pre-existing effect on brain structure^16-18^, which may or may not interact with stroke in determining post-stroke cognitive status. The finding that diabetes mellitus and smoking were associated with cognitive performance at both 3 months and 1 year, but not with change in cognitive scores, is consistent with these risk factors having a damaging effect on brain structure and cognition that is independent of the index stroke lesion. Fazekas score, diabetes mellitus and age were all associated with executive function in a multivariable ANCOVA model. This pattern of association is consistent with cerebral small vessel disease as a common factor underpinning these associations^19^.

Multiple cognitive domains can be affected by stroke ^20^, with the overall pattern of involvement varying from person to person. In addition to the MoCA score, which is used as a general index of cognition, scores of verbal episodic memory and executive function were included in this study. In cross-sectional analyses, age and smoking were associated with both verbal memory and executive function. Infarct volume also correlated with measures of verbal episodic memory, executive function and total MoCA score. These associations, common to more than one domain, were not sustained once other variables were taken into account, which is consistent with a previous study ^3^. The domain-specific association between lesion location and verbal memory, however, was found to be significant independent of other factors. Previous studies have shown a greater prevalence of cognitive impairment after left than right hemisphere stroke, and an association with infarction involving the cortex ^3^. So associations between cognitive impairment and lesion topography have been identified, albeit based on a crude schema to represent location. Collectively, the findings suggest that the profile of cognitive impairment in any individual is determined by a combination of pre-existing factors and the influence of particular lesion locations on specific cognitive networks and domains, accounting for the heterogeneity at an individual level in post-stroke cognitive profile.

The mechanisms that underlie the association between baseline IHD and *change* in cognitive scores are unclear. Recurrent embolization to the brain seems unlikely as the association was with IHD and not atrial fibrillation or heart failure. The association was specifically with coronary artery disease: participants were defined as having IHD if they had a history of an acute coronary syndrome or coronary revascularisation. The fourth Auckland Stroke Regional Outcomes Study (ARCOS-IV) reported that a history of coronary heart disease was associated with outcome in univariate analysis, although not when other socio-demographic factors were taken into account ^21^. The results are aligned with an increasing appreciation of the importance of the heart-brain axis for brain health. Future research should investigate those features of cardiac function that might underlie this association with IHD. Although we found no association with a diagnosis of AF, unrecognised AF or altered left ventricular function^18^ could contribute to compromise of the cerebral circulation. An alternative explanation is that IHD limits exercise tolerance and thereby interferes with participation in rehabilitation and likely beneficial effects of exercise intensity on neural plasticity^22^.

The strengths of the present study include a relatively large and well-phenotyped clinical cohort. The design allowed risk and demographic factors to be considered in parallel with infarct size and location, leading to an understanding of independent associations unlikely to arise from confounding. In addition, the use of specific tests of episodic memory and executive function, in addition to the widely used MoCA, made it possible to contrast the pattern of associations for general cognitive function and specific domains, providing evidence that some associations are domain-specific. Another strength is the use of continuous rather than binary cognitive outcomes (e.g. dementia). Also, unlike some comparable studies, we report associations with performance, and change in performance over time, separately, which has led to the observation that the factors associated with cross-sectional performance and longitudinal change are different. There are also weaknesses. The recruitment of patients with mild to moderate stroke without overt aphasia places limits on the generalisability of the findings. Lack of insight into pre-stroke cognitive function is a limitation common to many hospital-based cohort studies. Although symptomatic recurrent stroke was excluded by review of clinical histories and medical records, new asymptomatic infarcts were not excluded by serial MRI. It is likely that larger sample sizes would identify other factors associated with change in cognitive scores – for example, the association with T2DM was of marginal significance in this sample (no significant association with change in MoCA score, p=0.032 in multivariable ANCOVA).

Even with recent advances in reperfusion therapies in stroke, up to 40% of stroke survivors are left with at least moderate disability. Therapeutic innovation to support stroke recovery remains important. The present study raises the question of whether intervention targeted at the heart might play a role in supporting stroke recovery. Another potential implication is optimisation of the design of recovery trials based on knowledge of the factors that influence the natural history of recovery^5,23^.

## Data Availability

All data is available for sharing with qualified investigators and will be shared, in anonymized form, upon request, in accordance with the terms of the study ethical approvals.

